# Regional cardiac ^31^P-MRSI at 7T: detection of spatially heterogeneous patterns of myocardial energetic impairment

**DOI:** 10.64898/2026.09.04.26362251

**Authors:** Jabrane Karkouri, Will Watson, Tracy Horn, Marion Hill, Dennis Klomp, Michael Mallouppas, Catriona Bhagra, Stephen Hoole, Christopher T. Rodgers

## Abstract

**Background:** The phosphocreatine-to-ATP ratio (PCr/ATP), measurable by ³¹P magnetic resonance spectroscopic imaging (³¹P-MRSI), is a sensitive marker of myocardial energetic reserve. Regional energetic-mechanical coupling, evaluable only per-segment to avoid confounding from spatially heterogeneous disease, represents an important target for cardiac metabolic trials. We extend our previously validated 7T regional ³¹P-MRSI approach to patients with cardiovascular disease.

**Methods:** Twenty-one participants underwent same-day 7T ³¹P-MRSI and 3T CMR: 9 healthy volunteers, 4 with type 2 diabetes, 5 with heart failure, and 3 with ischaemic cardiomyopathy. PCr/ATP was mapped across six mid-ventricular AHA segments. Ejection fraction, global longitudinal strain, peak filling rate, and regional circumferential and radial strain were derived from 3T CMR. Group differences were assessed by Kruskal-Wallis test with Bonferroni-corrected pairwise comparisons; segment-level associations by per-segment Spearman correlation and linear mixed effects models.

**Results:** PCr/ATP differed significantly across groups (p = 0.005, η² = 0.58), ranging from 1.78 ± 0.21 in healthy volunteers to 1.10 ± 0.28 in heart failure. Mean PCr/ATP correlated with ejection fraction (rs = 0.65, p = 0.002), global longitudinal strain (rs = −0.63, p < 0.001), and peak filling rate (rs = 0.52, p = 0.023). Segment-level energetic-mechanical coupling was significant in the anteroseptal and inferoseptal segments (circumferential strain rs = −0.49 to −0.56; radial strain rs = 0.43 to 0.57, all p < 0.05), with no significant associations in other segments, consistent with a displacement-related measurement quality gradient. In ischaemic patients, PCr/ATP was significantly lower in infarct segments (0.79 ± 0.20) than remote myocardium (1.33 ± 0.22, p = 0.008, r = 0.62). All three ischaemic patients demonstrated remote PCr/ATP below the healthy volunteer median, with two below the 5th percentile (rank-sum p = 0.018; Fisher p = 0.045).

**Conclusions:** Regional 7T ³¹P-MRSI detects segment-level energetic differences across cardiovascular disease states and reveals spatially specific energetic-mechanical coupling confined to the septal segments in the current ungated acquisition. Energetic impairment in ischaemic cardiomyopathy extends into structurally preserved remote myocardium. These findings support larger studies using regional ³¹P-MRSI to investigate regional energetic-mechanical coupling as a target for metabolic cardiac therapies.

## Introduction

Myocardial phosphocreatine (PCr) buffers the continuous energy demands of the beating heart by rapidly regenerating adenosine triphosphate (ATP) via the creatine kinase reaction(1,2). The PCr/ATP ratio, measurable non-invasively by phosphorus-31 magnetic resonance spectroscopy (³¹P-MRS), has emerged as a sensitive in vivo marker of myocardial energetic reserve that is reduced across a broad spectrum of cardiovascular disease including heart failure, diabetic cardiomyopathy, hypertrophic cardiomyopathy, and ischaemic heart disease (3–14). Critically, energetic impairment often precedes overt contractile dysfunction, raising the prospect of PCr/ATP as an early biomarker of disease progression and a surrogate endpoint for therapeutic intervention (6,11).

Cardiac ³¹P-MRS has most commonly been performed by acquiring a single mid-septal voxel and taking it as representative of whole-heart energetics (6,11,15–19). The septal location is not arbitrary: reproducibility studies have demonstrated that the interventricular septum yields the most reliable and repeatable PCr/ATP measurements, likely because it is mechanically constrained, undergoes less through-plane displacement than the lateral and anterior walls, and consistently contains a mixture of myocardium and blood pool whose relative contributions can be corrected for(16,18). This technical rationale for septal acquisition carries an implicit assumption, however — that energetic changes in cardiovascular disease affect the whole left ventricle uniformly, such that a single well-chosen location is representative. For diffuse conditions such as dilated cardiomyopathy this assumption may be reasonable, but for diseases characterised by regional heterogeneity of metabolic impairment it is unlikely to hold. In ischaemic heart disease, energetic deficits are spatially confined to the infarct territory and potentially to a surrounding zone of hibernating or stunned myocardium, whilst remote segments may retain near-normal energetics(20,21). Averaging across the whole ventricle inevitably dilutes such focal deficits, masking their detection and precluding co-localisation with complementary regional functional metrics such as segmental strain. More broadly, if disease-related metabolic changes are spatially heterogeneous and a global acquisition spatially averages over affected and unaffected regions, a substantial portion of the statistical power to detect small energetic differences is lost — a critical limitation for the use of ³¹P-MRS as a clinical trial endpoint.

Advances in chemical shift imaging and spectroscopic imaging sequences, combined with the near-quadratic SNR gain at ultra-high field 7T — demonstrated as a 2.5-fold improvement in PCr SNR relative to 3T alongside a 45% reduction in quantification uncertainty(17,22) — have made regional multi-voxel ³¹P-MRS of the human heart technically feasible, enabling PCr/ATP mapping across segments aligned to the standard AHA myocardial segmentation scheme (23). Per-region cardiac ³¹P-MRS was first demonstrated in healthy volunteers at 2T (24)and 3T (18).

In light of the excellent whole-heart SNR obtained from 7T and advanced RF coils(25,26) in volunteers, we now test readiness to apply this approach in cardiac patients. Specifically: first, to verify that our regional acquisition detects the expected PCr/ATP reductions across patient groups — confirming that the measurements are reliable and not simply reporting a constant value irrespective of disease state — and to assess the extent to which the anteroseptal voxel is representative of whole-heart energetics across the left ventricle, directly testing the assumption that has underpinned the field for decades. Second, to use the wider range of PCr/ATP values available across a mixed disease cohort to extend the statistical leverage for interrogating the relationship between regional energetics and regional contractile function, building on our preliminary healthy volunteer findings(25). Third, to demonstrate that regional ³¹P-MRSI can directly resolve the energetic heterogeneity between infarcted and remote myocardium within individual ischaemic patients, and to characterise the energetic status of remote myocardium that appears structurally preserved on conventional imaging. The spectral quantification and segmental PCr/ATP mapping pipeline used in this study is made openly available alongside this manuscript.

## Methods

### Participants and Study Population

This was an observational study recruiting participants between late 2023 and early 2026 across three clinical sites: Royal Papworth Hospital, Cambridge University Hospitals NHS Trust, and Bedford Hospital NHS Trust. All 7T and 3T imaging was performed at the Wolfson Brain Imaging Centre, University of Cambridge. A total of 21 participants gave written consent and were recruited in accordance with the local Research Ethics Committee approval (Cambridge South Research Ethics Committee, REC: 23/EE/0048) across four groups: 9 healthy volunteers, 4 patients with type 2 diabetes mellitus, 5 patients with heart failure, and 3 patients with ischaemic cardiomyopathy. The healthy volunteers enrolled in this study were distinct from those reported in our prior feasibility work(25), and are reported here for the first time.

Patients with heart failure were recruited prior to SGLT-2 inhibitor therapy, and a clinical diagnosis of heart failure confirmed by prior imaging. HFrEF was defined as ejection fraction below 40% and HFpEF above 40%, by echocardiography, MRI, or another imaging modality. Diabetic patients required a diagnosis of type 2 diabetes mellitus without heart failure, with normal natriuretic peptide levels or elevated natriuretic peptides with normal cardiac function on echocardiogram. Healthy volunteers had no significant cardiovascular disease and no diagnosis of diabetes mellitus. Ischaemic patients were recruited from Royal Papworth Hospital following myocardial infarction and clinical stabilisation; heart failure and diabetic patients from outpatient cardiology and diabetology clinics; healthy volunteers from the local community. Exclusion criteria included factors preventing informed consent or MRI contraindications including pacemakers, cochlear implants, non-MR compatible implants, and claustrophobia.

Baseline characteristics are presented in Table 1. The cohort was deliberately designed to include a wide range of ages and disease severities, from young healthy volunteers (mean age 38 ± 11 years) to older patients with established cardiovascular disease (53–71 years across disease groups), in order to maximise statistical leverage for the regression analyses of energetics against functional metrics. Groups differed significantly in age (p = 0.002), GLS (p = 0.020), and PCr/ATP (p = 0.005). BMI and LVEF did not differ significantly across groups.

**Table 1:** Baseline characteristics of the study population. Data shown as mean ± SD. The Kruskal-Wallis test is a non-parametric test of whether the groups differ. Significant difference between the groups for that parameter is considered when P<0.05.

| Group | n | Sex (M/F) | Age | BMI | LVEF | GLS | PCr/ATP |
| --- | --- | --- | --- | --- | --- | --- | --- |
| Healthy control | 9 | 6/3 | 38.0 ± 11.2 | 24.6 ± 3.2 | 60.9 ± 6.1 | -17.2 ± 1.3 | 1.50 ± 0.28 |
| diabetic | 4 | 2/2 | 53.5 ± 10.5 | 26.9 ± 3.8 | 56.5 ± 5.4 | -16.9 ± 2.1 | 1.29 ± 0.28 |
| heart failure | 5 | 2/3 | 71.2 ± 8.2 | 27.7 ± 4.0 | 52.0 ± 9.8 | -14.0 ± 3.2 | 1.34 ± 0.46 |
| ischemic | 3 | 2/1 | 71.0 ± 4.6 | 35.1 ± 2.5 | 48.0 ± 11.1 | -11.5 ± 3.1 | 1.16 ± 0.17 |
| *p-value* |  |  | *0.002* | *0.044* | *0.150* | *0.021* | *0.376* |

*Data shown as mean ± SD. Sex shown as male/female. p-values from Kruskal-Wallis test.*

### 7T ^31^P-MRSI Measurements

#### Hardware

Scans were performed using a 7T MRI (Magnetom Terra, Siemens Healthcare, Erlangen, Germany) using a transmit/receive dipole-loop array comprising 8Tx/Rx ^1^H dipole elements, 8Tx/Rx ^31^P dipole elements, and 16 ^31^P Rx loops (Tesla Dynamic Coils, Zaltbommel, Netherlands) (27).

#### In vivo data acquisition

In vivo ³¹P-MRSI data were acquired as previously described(25). Briefly, we used a 3D CSI sequence with a 1s TR and 500 V_rms_ amplitude, selected to approximate the Ernst flip angle for PCr (T_1_ = 3.05 s, Ernst angle = 44° at 1 s TR) (22), 20×20×12 matrix and 30×35×30 cm³ FOV, yielding a nominal voxel size of 1.5×1.75×2.5 cm³, 2 averages at k=0 with acquisition weighting, Hamming filtering, 5kHz bandwidth and 1024 spectral points, and B1-insensitive train to obliterate signal (BISTRO) (28) saturation bands of 25mm thickness over the anterior chest wall and posterior back muscles. The total acquisition time was 30min.

The vendor tune-up B_0_ shim was used, as previously described (16,22,25). Respiratory and cardiac gating were not employed (29). The CSI grid was positioned on the mid-short axis localiser slice, with in-plane rotation and translation applied to maximise the number of voxels overlying the left ventricular myocardium.

#### Data Analysis

**Figure 1:**
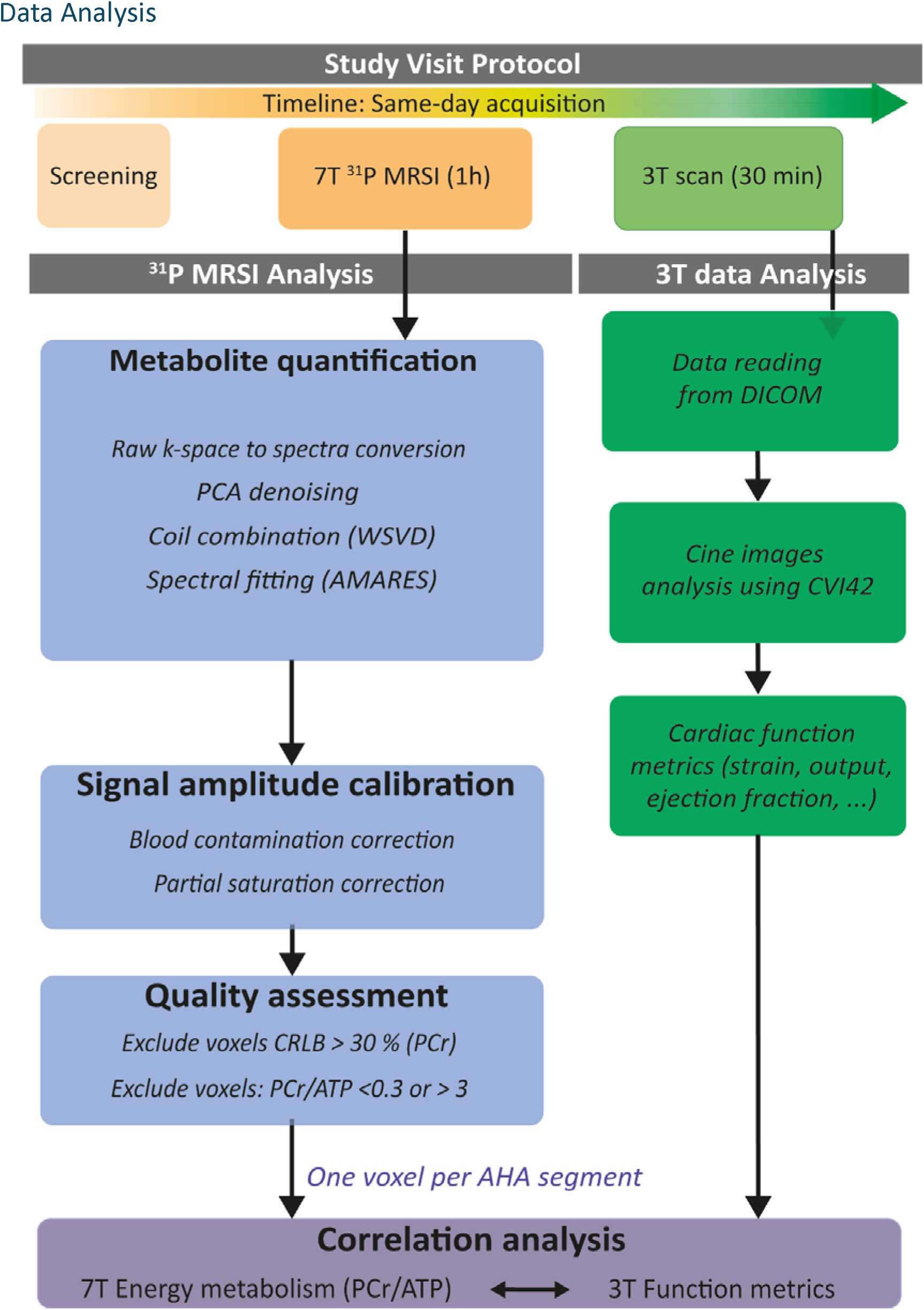
Study visit protocol and data analysis pipeline. All participants underwent same-day 7T ³¹P-MRSI (approximately 1 hour) and 3T CMR (approximately 30 minutes) at the Wolfson Brain Imaging Centre, Cambridge. Left panel (blue): ³¹P-MRSI processing pipeline, comprising raw k-space to spectra conversion, PCA denoising, coil combination using Whitened Singular Value Decomposition (WSVD), spectral fitting using the AMARES algorithm, blood contamination and partial saturation correction, and voxel-level quality assessment. Right panel (green): 3T CMR analysis pipeline, comprising DICOM data import and cine image analysis using CVI42 to derive cardiac functional metrics including strain, ejection fraction, and cardiac output. Metabolic and functional data were subsequently co-registered to AHA mid-ventricular segments for correlation analysis (purple).

Data were processed in MATLAB (Matlab R2023a, Mathworks Inc, Natick, MA, USA) using an updated version of the Open-Source Spectroscopy Analysis (OXSA) toolbox, adapted to support the Siemens 7T Terra MRI scanner (30). Principal component analysis (PCA)-based denoising was applied to the multi-channel image-space data (31). Signals from the different receive channels were combined using Whitened Singular Value Decomposition (WSVD) (32). Spectra were fitted using the Advanced Method for Accurate, Robust and Efficient Spectral fitting (AMARES) algorithm, with prior knowledge comprising 11 Lorentzian peaks encompassing PCr, γ-ATP, α-ATP, β-ATP, inorganic phosphate (Pi), diphosphoglycerate (2,3-DPG), phosphodiesters (PDE), and phosphomonoesters (PME).

Spectral fitting was performed on voxels selected for further analysis, localised to the myocardium only, and identified by overlay of the CSI grid on the anatomical localiser images. For quantitative comparison with 3T functional metrics, six voxels per subject were selected at standardised anatomical locations on the mid-interventricular short-axis slice, corresponding to the anterior, antero-septal, inferoseptal, inferior, inferolateral and anterolateral wall segments of the AHA mid-ventricular tier (23).

Blood- and saturation-corrected PCr/ATP ratios were calculated using literature T_1_ values of 3.05 s for PCr and 1.82 s for γATP (16,22), and flip-angles computed from phantom B1⁺ maps, as previously described (25).

Voxels with Cramér-Rao Lower Bounds (CRLBs) for PCr amplitude exceeding 30% were excluded. Voxels with PCr/ATP <0.3 or PCr/ATP > 3.0 were also excluded as these values were considered physiologically implausible.

#### Code Availability

The ³¹P-MRSI analysis code, including the adapted OXSA toolbox and AHA segment assignment pipeline, is publicly available at [GitHub/GitLab repository URL to be inserted upon acceptance]. The anonymised participant data is available from the senior author upon reasonable request, subject to institutional data governance approval and a formal data transfer agreement.

### 3T-CMR

Cardiac function was assessed using 3T MRI (Prisma, Siemens, Erlangen, Germany) on the same day as the 7T ³¹P-MRSI scan. Scans included localisers in three orthogonal planes, followed by two-chamber, four-chamber, and short-axis balanced steady-state free precession (bSSFP) cines (BEAT_Cine_TruFISP) (33–35).

Left ventricular volumes, ejection fraction, and cardiac mass were quantified from short-axis cine stacks using CVI42 (Circle Cardiovascular Imaging, Calgary, Canada) by manual contouring of endo- and epicardial borders at end-diastole and end-systole. Global longitudinal strain and peak filling rate were derived from the long and short-axis cine acquisitions. Regional circumferential and radial strain were assessed from the mid-ventricular short-axis slice, with the left ventricle divided into six segments according to the AHA 17-segment model(23): anterior, anteroseptal, inferoseptal, inferior, inferolateral, and anterolateral. For each participant, ³¹P-MRSI voxels were assigned to the corresponding mid-ventricular AHA segments based on anatomical overlay of the CSI grid on the short-axis localiser.

### Statistical analysis

Statistical analyses were performed in MATLAB (R2023a, MathWorks Inc., Natick, MA, USA). Given the small sample sizes, non-parametric methods were used throughout unless otherwise specified. A two-sided significance threshold of p < 0.05 was applied, and data are presented as mean ± standard deviation unless otherwise stated. Effect sizes are reported as eta-squared (η²) for the Kruskal-Wallis test, rank-biserial correlation (r) for Wilcoxon rank-sum tests, and marginal R² for linear mixed effects models(36). Spearman rs values are reported directly as measures of association strength. Effect size benchmarks follow established conventions(37).

First, we aimed to verify detection of expected group-level energetic differences. Group differences in PCr/ATP and clinical variables were assessed using the Kruskal-Wallis test. Where a significant overall group effect was detected, targeted pairwise comparisons between healthy volunteers and each disease group were performed using the Wilcoxon rank-sum test with Bonferroni correction for three comparisons (adjusted threshold p < 0.017), as these comparisons were specified a priori as the primary scientific question.

Second, we aimed to assess whether the anteroseptal voxel is representative of whole-heart energetics. The correlation between anteroseptal PCr/ATP and whole-myocardium mean PCr/ATP was assessed using Spearman rank correlation across all cohorts. A forced-through-origin ordinary least squares regression (y = ax) was fitted to characterise the relationship relative to the line of identity (y = x), with a bootstrap 95% confidence interval on the slope used to assess whether the anteroseptal voxel introduces systematic bias.

Third, we investigated the energetic-functional associations. To test whether energetic-functional associations are detectable at the whole-heart level, Spearman rank correlation was applied between per-participant mean PCr/ATP and global functional metrics (LVEF, GLS, and peak filling rate). For segment-level associations between regional PCr/ATP and myocardial strain, per-segment Spearman correlations were computed separately for each of the six AHA segments. Linear mixed effects models with patient as a random intercept were additionally fitted to the pooled segment-level data to account for the non-independence of multiple segments within the same individual, and to assess whether any pooled association survived adjustment for age and BMI as fixed covariates.

Finally, we investigated regional energetic heterogeneity in ischaemic myocardium. Regional differences in PCr/ATP between infarct and remote segments in ischaemic patients were assessed using the Wilcoxon rank-sum test, with infarct segments defined according to subject-specific coronary territory mapping. To assess whether remote myocardium falls within the healthy reference range, mean PCr/ATP across non-infarcted segments per patient was compared to the healthy volunteer distribution using the Wilcoxon rank-sum test. Fisher’s exact test was used to compare the proportion of ischaemic patients falling below the healthy volunteer 5th percentile against the proportion of healthy volunteers below this threshold. Given the sample size of n = 3 ischaemic patients, these comparisons are descriptive in nature and interpreted alongside effect sizes rather than p-values alone.

## Results

### PCr/ATP maps and spectral quality

Representative PCr/ATP maps overlaid on mid-ventricular short-axis anatomical images are shown for one participant from each group in Figure 2. In the healthy control (panel A, mean PCr/ATP = 1.83, between-segment SD = 0.35), PCr/ATP is relatively uniform across all left ventricular wall segments, with consistent values throughout the myocardium. In the type 2 diabetic patient (panel B, mean PCr/ATP = 1.67), PCr/ATP is modestly and uniformly reduced relative to the healthy control, with all segments similarly affected. In the heart failure patient (panel D, mean PCr/ATP = 0.70), PCr/ATP is globally and severely reduced across all visible myocardial segments. In the ischaemic patient (panel C, mean PCr/ATP = 1.33), a focal reduction in PCr/ATP is visible in the inferolateral wall corresponding to the infarct territory, whilst the remote myocardium shows relatively preserved values — illustrating the regional heterogeneity that global single-voxel acquisitions would average over. These representative maps demonstrate the dynamic range of myocardial PCr/ATP detectable by 7T ³¹P-MRSI across disease states. Representative spectra of good quality for each cohort are shown in Supplementary Figures SI1–4

**Figure 2:**
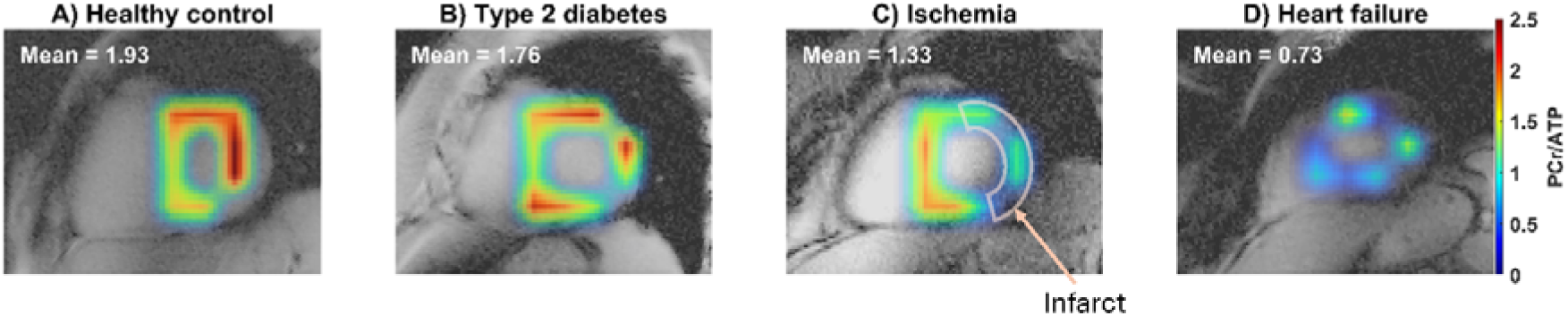
Representative PCr/ATP ratio maps overlaid on mid-ventricular short-axis anatomical images for one participant from each group: healthy control (A), type 2 diabetes mellitus (B), ischaemic cardiomyopathy with a inferolateral infarct territory (C), and a single heart failure patient prior to (D). Mean PCr/ATP values for the displayed participant are shown in each panel. The colour scale is common to all panels.

### Group differences in myocardial energetics

Per-participant mean PCr/ATP ratios across the six AHA segments are shown in Figure 3. Values differed significantly between groups (Kruskal-Wallis p = 0.005, η² = 0.58), with the large effect size indicating that group membership accounts for a substantial proportion of the variance in PCr/ATP across the cohort. Healthy volunteers demonstrated the highest mean PCr/ATP (1.78 ± 0.21), followed by diabetic patients (1.49 ± 0.31), ischaemic patients (1.12 ± 0.16), and heart failure patients (1.10 ± 0.28). Targeted pairwise comparisons between healthy volunteers and each disease group, with Bonferroni correction for three comparisons (adjusted threshold p < 0.017), revealed significant reductions in heart failure (p = 0.002, r = 0.96) and ischaemic patients (p = 0.009, r = 1.00) relative to healthy volunteers. The comparison between healthy volunteers and diabetic patients did not reach significance after correction (p = 0.148), though the moderate effect size (r = 0.56) is consistent with a true energetic reduction that the present sample size was insufficient to confirm statistically. Heart failure and ischaemic patients did not differ significantly from each other (p = 0.571), consistent with similar degrees of global energetic impairment in both conditions.

**Figure 3:**
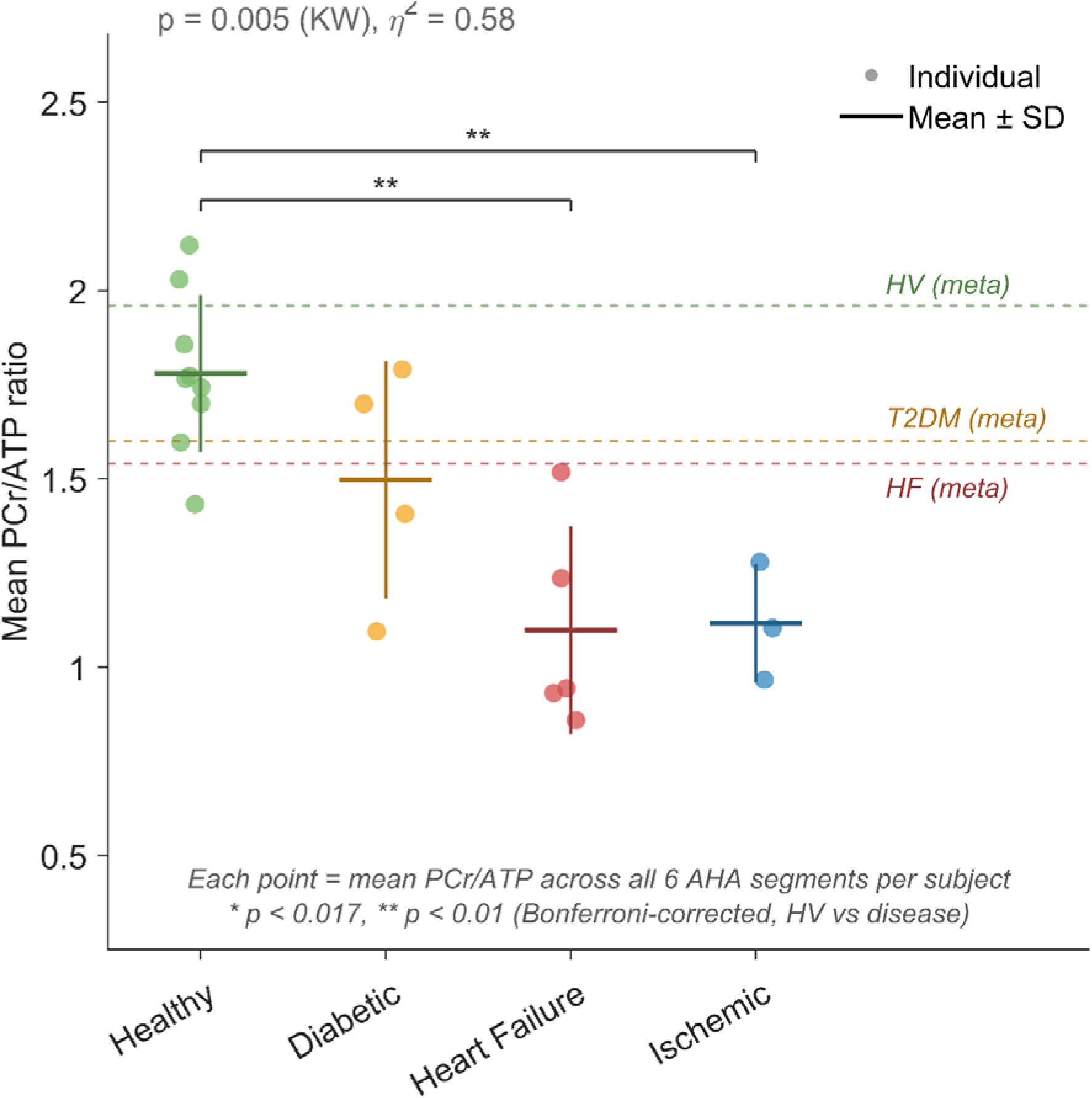
Group differences in myocardial PCr/ATP ratio. Each point represents one participant’s mean PCr/ATP averaged across all available mid-ventricular AHA segments. Horizontal bars indicate group mean ± SD. Groups differed significantly by Kruskal-Wallis test (p = 0.005, η² = 0.58). Pairwise comparisons between healthy volunteers and each disease group used the Wilcoxon rank-sum test with Bonferroni correction for three comparisons (* p < 0.01, adjusted threshold p < 0.017). HF = heart failure.

### Correlations between myocardial energetics and cardiac function

Spearman rank correlations between mean PCr/ATP and global functional parameters are shown in Figure 4A–C. Significant positive correlations were observed with LVEF (rs = 0.65, p = 0.002) and peak filling rate (rs = 0.52, p = 0.023), and a significant negative correlation with GLS (rs = −0.63, p = 0.002), indicating that higher PCr/ATP was associated with better preserved systolic and diastolic function across the cohort. The association with GLS was the strongest of the three, consistent with GLS being a sensitive marker of subclinical myocardial dysfunction that may be more directly coupled to myocardial energetic demand than ejection fraction alone. After adjusting for age and BMI, PCr/ATP remained independently associated with both LVEF and GLS, and adding these covariates did not significantly improve model fit, confirming that the global energetic-functional associations are not driven by the age difference between cohorts.

**Figure 4:**
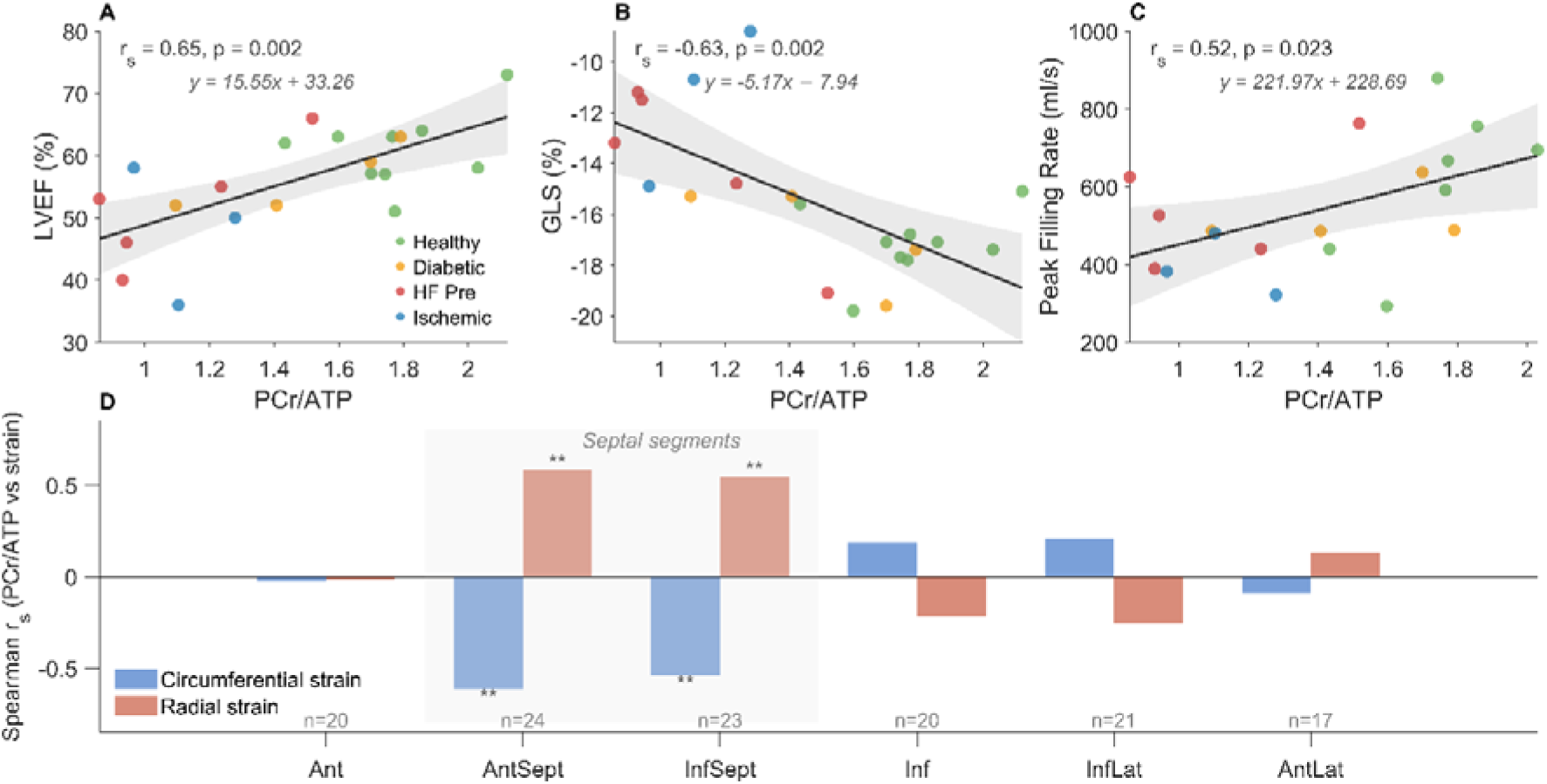
Correlations between myocardial PCr/ATP and cardiac functional parameters. (A–C) Spearman rank correlations between per-participant mean PCr/ATP and global functional parameters: (A) left ventricular ejection fraction (LVEF), (B) global longitudinal strain (GLS), and (C) peak filling rate. Each point represents one participant, coloured by disease group. The regression line with 95% confidence band is shown in grey; the regression equation (y = βx + β₀) and Spearman rs and p-values are displayed within each panel. (D) Per-segment Spearman rank correlations between regional PCr/ATP and circumferential strain (blue) and radial strain (red) across the six mid-ventricular AHA segments. Bar height represents the Spearman rs for each segment; significant associations are marked ( p < 0.05, ** p < 0.01). Grey shading indicates the anteroseptal and inferoseptal segments where significant coupling was observed. Sample sizes per segment are shown below the x-axis. PCr, phosphocreatine; ATP, adenosine triphosphate; LVEF, left ventricular ejection fraction; GLS, global longitudinal strain; AHA, American Heart Association; rs, Spearman rank correlation coefficient.“

At the segment level, linear mixed effects models with patient as a random intercept revealed a significant association between regional PCr/ATP and circumferential strain (β = −2.13, p = 0.040, R =0.19), and a non-significant trend for radial strain (β = 4.54, p = 0.171, R =0.23). Per-segment Spearman correlations revealed that the circumferential strain association was driven primarily by the septal segments — anteroseptal (CS: rs = −0.49, p = 0.024; RS: rs = 0.43, p = 0.049) and inferoseptal (CS: rs = −0.56, p = 0.010; RS: rs = 0.57, p = 0.010) — with no significant associations observed in the anterior, inferior, inferolateral, or anterolateral segments (all p > 0.15, Figure 4D). Both associations were abolished when segment identity was included as an additional random effect (CS: β = 0.03, p = 0.961; RS: β = −0.79, p = 0.718), confirming that the energetic-mechanical coupling is segment-specific rather than globally distributed.

### Regional Distribution of Myocardial PCr/ATP Ratio

Regional PCr/ATP ratios across the six AHA segments are shown in Figure 6. In healthy volunteers, PCr/ATP values were broadly consistent across segments, with a modest relative reduction at the anteroseptal and inferoseptal segments compared to the anterior and anterolateral walls (Figure 5A). This pattern was consistent across individual subjects, and the narrow 95% CI of the healthy volunteer profile reflects the relatively low between-subject variability in regional PCr/ATP in the absence of disease. Notably, PCr/ATP values in all disease cohorts fell below the healthy volunteer reference band across every segment, with heart failure patients showing the most pronounced and spatially consistent reduction. In ischaemic patients, represented by remote non-infarcted myocardium only, values fell within or just below the healthy reference corridor at the group level, whilst the infarcted segments showed markedly lower PCr/ATP, directly visualising the focal energetic deficit. The regional profiles of the diabetic and heart failure cohorts were spatially homogeneous — all segments depressed to a similar degree relative to healthy volunteers — in contrast to the ischaemic cohort where the infarct territory was selectively and severely impaired. This spatial pattern, with uniform depression in diffuse disease and focal depression in ischaemia, is consistent with the underlying pathophysiology of each condition and supports the validity of the regional acquisition.

**Figure 5:**
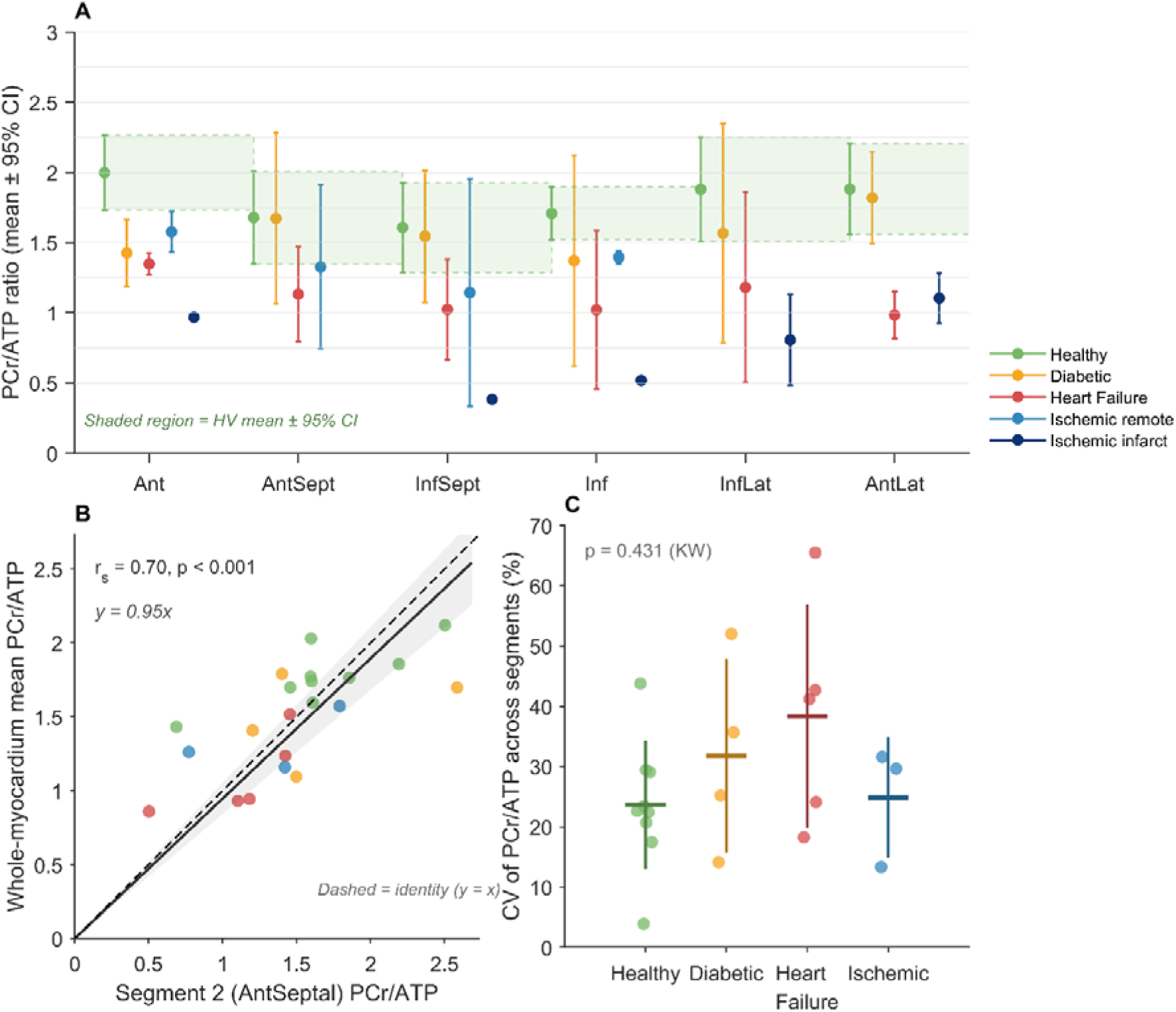
Regional myocardial PCr/ATP distribution across healthy volunteers and patient cohorts. (A) PCr/ATP ratio across the six mid-ventricular AHA segments for each cohort. Points and error bars represent cohort mean ± 95% CI, with a small horizontal offset applied per group for clarity. The green shaded region and dashed lines represent the 95% CI of the healthy volunteer mean profile. Ischaemic remote and infarct segments are shown separately as light and dark blue respectively. (B) Forced-through-origin regression of anteroseptal (Segment 2) PCr/ATP against whole-myocardium mean PCr/ATP across all cohorts. The solid line shows the fitted regression (y = 0.95x) and the dashed line the identity (y = x). The regression slope was not significantly different from 1.0 (bootstrap 95% CI: [0.859, 1.062]), indicating that the anteroseptal voxel is an unbiased estimator of whole-myocardium energetic status on average. (C) Coefficient of variation (CV%) of PCr/ATP across available mid-ventricular segments per subject. Individual points represent subjects; horizontal bars indicate cohort mean ± SD. No significant difference in regional metabolic heterogeneity was observed between cohorts (Kruskal-Wallis p = 0.431).

**Figure 6:**
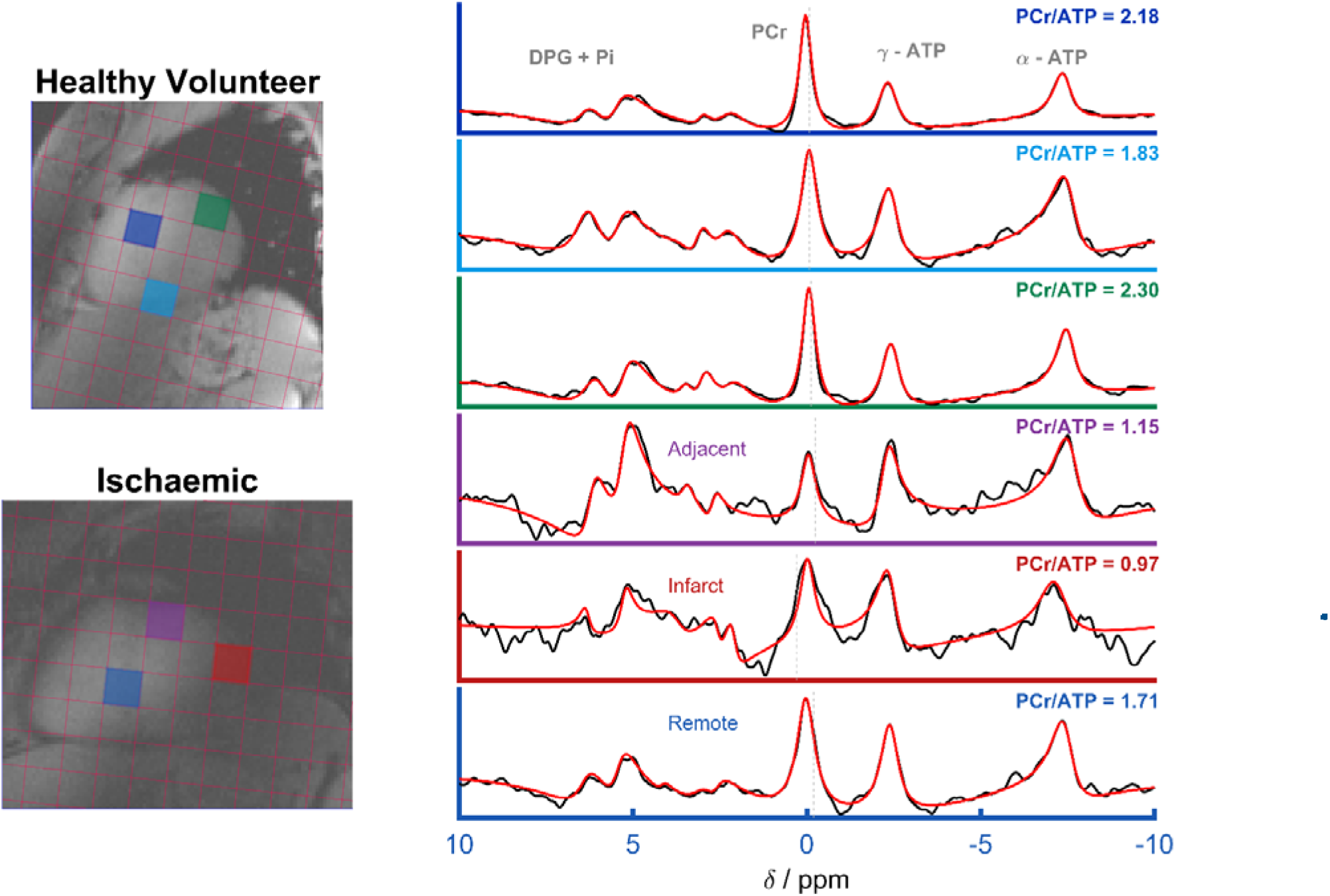
Representative ³¹P spectra from selected AHA segments in a healthy volunteer (top, three panels) and an ischaemic patient (bottom, three panels), with voxel locations indicated by colour-coded overlays on the corresponding mid-ventricular short-axis anatomical images. For the healthy volunteer, spectra are shown from three myocardial segments with PCr/ATP ratios of 2.18, 1.83, and 2.30, illustrating the regional variation in energetic status present in healthy myocardium. For the ischaemic patient, spectra are shown from an infarct segment (PCr/ATP = 0.97), an adjacent border zone segment (PCr/ATP = 1.15), and a remote segment (PCr/ATP = 1.71). The fitted model (red) is superimposed on the raw data (black) for each spectrum. Key metabolite peaks are labelled: phosphocreatine (PCr), γ-, α-ATP, and 2,3-diphosphoglycerate with inorganic phosphate (2,3-DPG + Pi). The progressive reduction in PCr amplitude relative to γATP from remote to border zone to infarct segments illustrates the spatial gradient of energetic impairment detectable by 7T ³¹P-MRSI.

The anteroseptal PCr/ATP ratio correlated significantly with whole-myocardium mean PCr/ATP across all cohorts (Spearman rs = 0.70, p < 0.001, Figure 5B). A forced-through-origin regression yielded a slope of 0.95 (bootstrap 95% CI: [0.859, 1.062]), which was not significantly different from the identity slope of 1.0, indicating that the anteroseptal voxel provides an essentially unbiased estimate of whole-myocardium mean PCr/ATP on average. The scatter around the regression line reflects genuine between-segment heterogeneity in individual patients that is lost when only a single voxel is acquired.

The coefficient of variation of PCr/ATP across myocardial segments did not differ significantly between cohorts (Kruskal-Wallis p = 0.431, Figure 5C), with mean CV of 23.6% in healthy volunteers, 31.7% in diabetic patients, 38.4% in heart failure patients, and 24.9% in ischaemic patients (remote segments only). For ischaemic patients, CV was computed across remote segments only, excluding the infarcted territory; the true whole-heart CV in this group would be expected to be substantially higher given the focal PCr/ATP reduction in the infarct zone.

### Regional energetic heterogeneity in ischaemic cardiomyopathy

Representative ³¹P spectra from a healthy volunteer and an ischaemic patient are shown in Figure 6, with voxel locations indicated by colour-coded overlays on the anatomical images. In the healthy volunteer, PCr/ATP ratios across three myocardial segments ranged from 1.83 to 2.30. In the ischaemic patient shown, a clear spatial gradient of energetic impairment was evident: the infarct segment showed a markedly reduced PCr/ATP of 0.97, the adjacent border zone an intermediate value of 1.15, and the remote segment a comparatively preserved ratio of 1.71. Across the three ischaemic patients, PCr/ATP was significantly lower in infarct segments compared to remote segments (Wilcoxon rank-sum p = 0.008, r = 0.62, Figure 6B), confirming that the regional energetic deficit observed in representative spectra is consistent across the cohort.

Mean PCr/ATP ratios across remote non-infarcted and infarcted myocardial segments in ischaemic patients are shown alongside the healthy volunteer distribution in Figure 7. A clear descending gradient was observed from healthy myocardium through ischaemic remote to ischaemic infarct segments. Mean remote PCr/ATP was significantly lower than in healthy volunteers (Wilcoxon rank-sum p = 0.018, r = −0.93), and mean infarct PCr/ATP was significantly lower than both healthy volunteers (p = 0.0091, r = −1.0) and remote segments (p = 0.008, r = 0.62). Two of three ischaemic patients demonstrated mean remote PCr/ATP values below the healthy volunteer 5th percentile; the proportion of ischaemic remote patients below this threshold was significantly greater than expected from the healthy volunteer distribution (Fisher’s exact test p = 0.045). These findings suggest that energetic impairment in subacute myocardial infarction is not confined to the infarcted territory but extends into structurally preserved remote myocardium.

**Figure 7:**
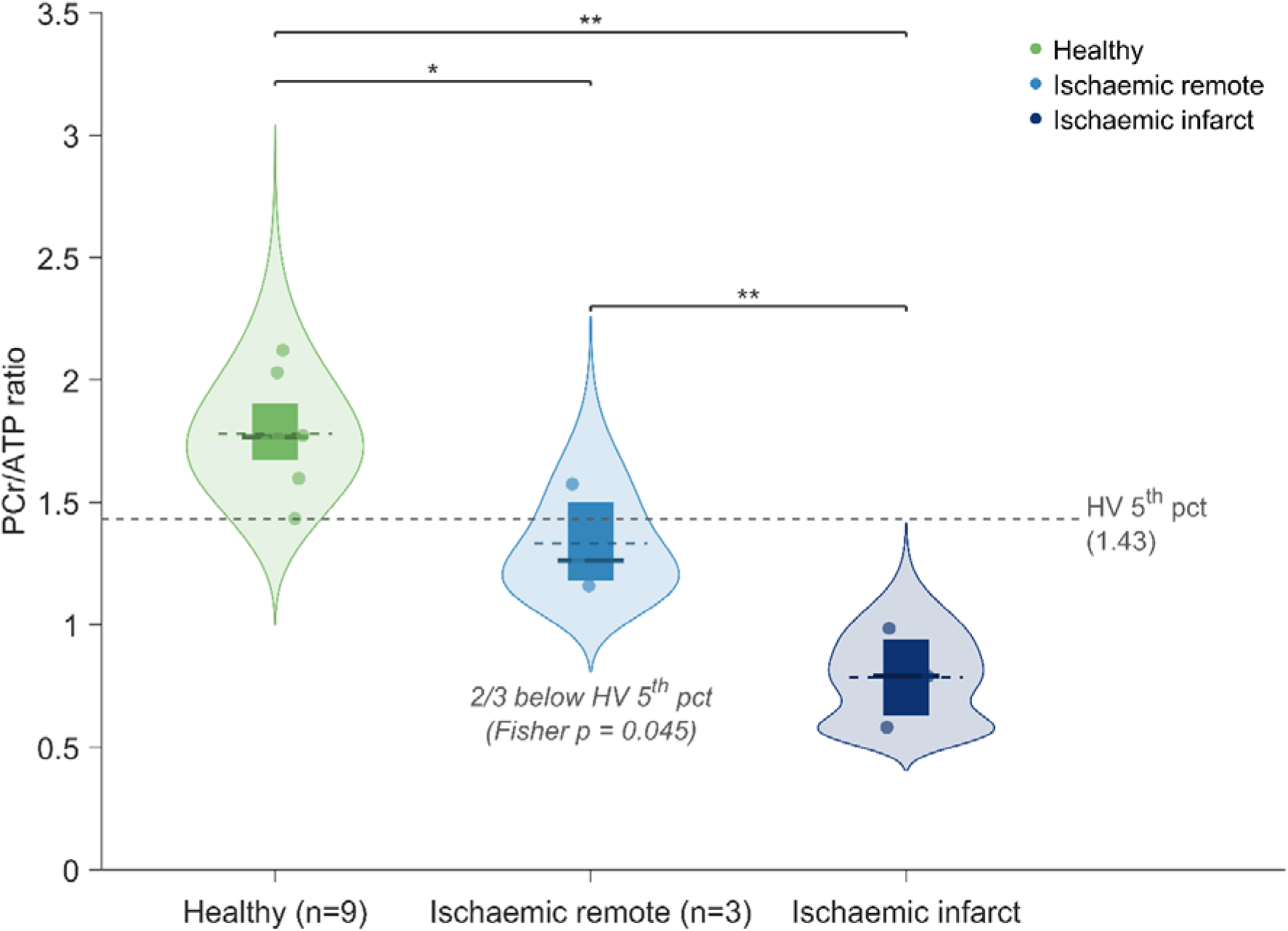
Mean PCr/ATP ratio across remote (non-infarcted) AHA segments in ischaemic patients (n=3, green dots) overlaid on the healthy volunteer distribution (n=X, blue). The dashed line indicates the 5th percentile of the healthy volunteer distribution. Individual patient identifiers are shown. The horizontal bar indicates the group mean.

## Discussion

### Group Differences in Myocardial Energetics

The observed pattern of PCr/ATP across groups is consistent with established cardiac ³¹P-MRS literature. Healthy controls showed mean PCr/ATP of 1.78 ± 0.21, consistent with values of 1.91 ± 0.36 reported by Ellis et al. at 7T, 1.85 ± 0.37 by Gosselink et al (26) and 1.79 ± 0.22 reported in our healthy volunteer cohort volunteers (16,22,25). Heart failure patients had a lower PCr/ATP of 1.09 ± 0.28, consistent with the well-established energetic deficit in heart failure and in keeping with values reported by Neubauer et al. at 3T (11), Weiss et al. (38) and Stoll et al. at 7T in dilated cardiomyopathy (17).

Diabetic patients had mean PCr/ATP of 1.49 ± 0.31 which is modestly reduced compared to healthy volunteers, consistent with Valkovič et al. who reported a similar relative reduction in type 2 diabetes at 7T (18% reduction compared to 16% in the present study) (39). The relatively preserved PCr/ATP in diabetics compared to heart failure patients is consistent with their being at an earlier stage of metabolic dysfunction.

### Correlations Between Myocardial Energetics and Cardiac Function

Spearman correlations between mean PCr/ATP and global functional parameters revealed significant associations with GLS (rs = −0.63, p = 0.002), LVEF (rs = 0.65, p = 0.002), and peak filling rate (rs = 0.52, p = 0.023). The strong correlation with GLS is noteworthy, as GLS is a sensitive marker of subclinical myocardial dysfunction that captures longitudinal fibre shortening and may be more directly coupled to myocardial energetic demand than ejection fraction. These findings are consistent with prior reports of PCr/ATP-function relationships at lower field strengths (8,12).

The association between PCr/ATP and circumferential strain is consistent with prior global-level observations in aortic stenosis, where reduced PCr/ATP was an independent predictor of peak circumferential strain (40), and with studies linking impaired myocardial energetics to subclinical contractile dysfunction in diabetic cardiomyopathy (41). We extended these observations to segment-level analysis across a mixed multi-disease cohort at 7T.

At the segment level, the energetic-mechanical associations were driven specifically by the anteroseptal and inferoseptal segments, with no significant coupling detected in the anterior, inferior, inferolateral, or anterolateral walls. This segment-specificity maps closely onto published measurements of regional cardiac displacement from short-axis CMR: epicardial displacement in the mid-ventricular tier ranges from 4.3 ± 0.8 mm in the inferoseptal segment to 5.5 ± 1.4 mm in the anterior wall in healthy controls, with the septal segments consistently showing the least through-plane motion across all cardiac disease states(42). In the absence of cardiac gating and prospective motion correction in the present acquisition, segments with greater through-plane displacement will experience more partial volume contamination per TR, increasing spectral noise and attenuating the detectable energetic-mechanical association. The septal segments, constrained by the interventricular septum and undergoing predominantly in-plane thickening, are relatively protected from this effect. The concordance between the segments showing significant PCr/ATP–strain coupling and those with least cardiac displacement therefore has two complementary interpretations: first, that genuine energetic-mechanical coupling is detectable where measurement quality is highest; and second, that improved motion correction — enabled by interleaved ¹H acquisition within the ³¹P sequence as demonstrated in our published sequence building block framework(43) — could recover these associations in the higher-motion segments in future studies. The marginal R² from the pooled model for circumferential strain (R² = 0.19) and the absence of a significant pooled association for radial strain (R² = 0.23, p = 0.171) are consistent with this interpretation: energetic-mechanical coupling is genuine but spatially heterogeneous, with detectable associations confined to the septal segments where spectral quality is highest, and attenuated to non-significance when averaged across all segments including the noisier lateral and inferior walls.

### Regional Energetic Heterogeneity and Methodological Validation

Regional PCr/ATP mapping across AHA segments revealed a consistent spatial pattern in healthy volunteers, with higher values in the anterior wall, a relative reduction at the anteroseptal and inferoseptal segments, and partial recovery toward the anterolateral wall. Whether this pattern reflects genuine regional metabolic heterogeneity heterogeneity with altered mechano-energetic coupling in more mobile segments or is partly attributable to spatially structured acquisition artefact — including residual B_1_⁺ inhomogeneity following phantom-derived correction, regional differences in cardiac motion, or partial volume effects from adjacent chest wall musculature — cannot be resolved with the current protocol, and prospective studies with subject-specific in vivo B_1_⁺ mapping are required to address this definitively.

The significant reduction in PCr/ATP in infarct segments relative to remote myocardium (p = 0.008), detectable across only three patients, directly validates the premise that regional ³¹P-MRSI can resolve spatially confined energetic deficits that would be masked by global voxel averaging. This is a limitation that was reflected in the relatively preserved global mean PCr/ATP of the ischaemic group, which obscured the true magnitude of infarct territory impairment. The magnitude of PCr/ATP reduction in infarct segments [0.79 ± 0.20 vs 1.33 ± 0.22 in remote segments] is consistent with prior 3T studies of ischaemic myocardium: Beer et al. reported PCr/ATP of 0.49 ± 0.23 in ischaemic versus 0.77 ± 0.17 in remote myocardium in post-MI patients, and Yabe et al. reported PCr/ATP of 0.94 ± 0.41 in coronary artery disease versus 1.80 ± 1.03 in controls (9,21). It also aligns with the mechanistic framework established in animal models by Friedrich et al. (44). The spatial selectivity demonstrated here at 7T goes beyond what was achievable at lower field strengths, where SNR constraints precluded AHA-aligned segment-level mapping.

All three ischaemic patients demonstrated mean remote PCr/ATP values below the healthy volunteer median, with two falling below the 5^th^ percentile of the healthy volunteer distribution. This is consistent with prior experimental and human evidence that energetic impairment after myocardial infarction extends beyond the infarcted territory: preclinical ³¹P chemical shift imaging has demonstrated that PCr decreases uniformly in non-infarcted remodelled myocardium independently of distance from the scar, with the magnitude of reduction inversely correlating with infarct size (44); the proposed mechanism involves tethering of the border zone to the infarct, increasing wall stress and energy demand in segments that are not themselves ischaemic (20); and in humans, reduced PCr/ATP in residual viable myocardium has been attributed to abnormal transmural distribution of high-energy phosphate compounds and to mismatch between oxygen delivery and demand (21). The clinical implications are potentially significant: the ability to distinguish energetically impaired infarct territory from metabolically compromised but structurally preserved remote myocardium could inform decisions around revascularisation, particularly in the context of hibernating myocardium. Larger studies with systematic correlation of regional PCr/ATP against late gadolinium enhancement extent and post-revascularisation functional recovery are needed to explore this hypothesis.

A key methodological concern in regional ³¹P-MRSI acquired without motion correction, B1 mapping, or cardiac gating is that observed regional PCr/ATP differences may partly reflect acquisition artefact rather than true metabolic heterogeneity, specifically that more mobile myocardial segments moving in and out of the acquisition voxel may yield artifactually different values compared to more static segments. Motion artefact could manifest in two distinct ways: as large systematic biases in absolute PCr/ATP values, or as increased measurement noise that attenuates the detectability of genuine associations without producing systematic offsets. If motion were the dominant source of large systematic biases, one would expect the coefficient of variation of PCr/ATP across segments to differ between cohorts with different motion characteristics. The absence of a significant difference in CV across cohorts (p = 0.431) argues against motion as a dominant driver of systematic absolute value biases. However, this does not exclude motion as a source of increased measurement noise in higher-displacement segments — consistent with the segment-specific pattern of energetic-functional associations described above. The correlation between anteroseptal PCr/ATP and whole-myocardium mean PCr/ATP was strong (r = 0.70, p < 0.001), confirming that a single septal acquisition captures meaningful information about global energetic status. A forced-through-origin regression yielded a slope of 0.95 (bootstrap 95% CI: [0.859, 1.062]), not significantly different from the identity slope of 1.0, confirming that the anteroseptal voxel provides an essentially unbiased estimate of whole-myocardium PCr/ATP on average. Prospective studies incorporating motion correction and B_1_ mapping will be required to fully characterise the contribution of acquisition artefact to regional PCr/ATP heterogeneity. It should additionally be noted that respiratory motion may contribute to regional PCr/ATP variation, particularly in segments that are more susceptible to diaphragmatic displacement, and the present analysis cannot fully separate cardiac from respiratory motion effects without respiratory gating or navigator data.

Apps et al. demonstrated absent bicarbonate signal in non-viable transmural infarct segments and preserved oxidative metabolism in viable subendocardial infarction using hyperpolarised ¹³C-pyruvate (45), showing that metabolic imaging can distinguish viable from non-viable myocardium. Our ³¹P-MRSI approach provides a complementary window into myocardial energetics that does not require hyperpolarisation infrastructure or exogenous tracer administration. However, it does require a 7T scanner and a dedicated cardiac ³¹P coil, representing a significant infrastructure investment, and the more restrictive implant safety policies at 7T relative to clinical field strengths remain an important practical consideration for patient accessibility.

### Limitations

We acknowledge the following limitations of our study. Cohort sizes are intentionally modest, as the primary aim was technical validation and proof-of-concept demonstration rather than definitive clinical inference. The absence of significance for some comparisons should therefore not be interpreted as evidence of no effect. Furthermore, the absence of an age-matched or older healthy control group means that the observed differences in PCr/ATP and functional metrics between healthy volunteers and disease cohorts cannot be fully disentangled from the effects of normal cardiac ageing, which independently reduces myocardial PCr/ATP by approximately 0.02 per year in healthy individuals(46). Inclusion of an older healthy cohort will be an important design consideration for future studies. Whilst the age-adjusted sensitivity analyses confirmed that the energetic-functional associations were not driven by age alone, a dedicated older healthy cohort would provide a more direct test of this. All acquisitions were performed at a single centre on a single 7T platform, and generalisability to other sites and scanner configurations remains to be established. The ³¹P-MRSI acquisition provides PCr/ATP averaged across relatively large voxels aligned to AHA segments, and partial volume effects from blood pool, epicardial fat, and adjacent tissue cannot be entirely excluded, although blood saturation correction was applied.

### Future Study Design

Power calculations based on the observed effect sizes and measurement repeatability from our prior reproducibility study (25) suggest that group-level PCr/ATP differences of the magnitude observed here would be detectable with approximately n = 6 patients per group at 80% power, confirming that adequately powered multi-group studies are achievable with modest cohort sizes — consistent with the significant pairwise differences detected here despite n = 3–5 per group. For segment-level energetic-mechanical associations, approximately 10 subjects would be required to detect septal coupling at 80% power with Bonferroni correction for six segment comparisons. In higher-motion lateral and inferior segments, the four-fold higher coefficient of repeatability (CR = 0.88 vs 0.22 in the septum) substantially attenuates the observable correlation, requiring approximately 25 subjects even if true biological coupling were equivalent — quantitatively confirming that prospective motion correction via interleaved ¹H acquisition (43) is a prerequisite for recovering energetic-mechanical associations across all segments. MATLAB code for these power calculations is provided in Supplementary Material.

## Conclusion

Regional cardiac ³¹P-MRSI at 7T is feasible across a spectrum of cardiovascular disease and demonstrates sufficient sensitivity to detect group-level energetic impairment, resolve intra-myocardial metabolic heterogeneity between infarcted and remote segments in ischaemic patients, and reveal spatially specific energetic-mechanical coupling in the septal myocardium. Energetic impairment in ischaemic cardiomyopathy extends beyond the infarct territory into structurally preserved remote myocardium — a finding with potential implications for risk stratification and therapy targeting. The single septal voxel, whilst an unbiased estimator of whole-heart energetics on average, cannot capture this regional heterogeneity. The spectral quantification and segmental PCr/ATP mapping pipeline, made openly available alongside this manuscript with test data and unit tests, provides a foundation for larger prospective studies of regional myocardial energetics across the cardiovascular disease spectrum.

## Supporting information

Supplementary code for power calculation

## Data Availability

The ^31^P-MRSI analysis code, including the adapted OXSA toolbox and AHA segment assignment pipeline, is publicly available at [GitHub/GitLab repository URL to be inserted upon acceptance]. The anonymised participant data is available from the senior author upon reasonable request, subject to institutional data governance approval and a formal data transfer agreement.

## Acknowledgements

This work was supported by the EU Horizon Europe project “MITI” #101058229. This research was supported by the NIHR Cambridge Biomedical Research Centre (NIHR203312) and the NIHR Applied Research Collaboration East of England. The views expressed are those of the author(s) and not necessarily those of the NIHR or the Department of Health and Social Care. The 7T MRI is supported by the Medical Research Council [MR/M009041/1; MR/M008983/1; UKRI2679; and UKRI790] and the Isaac Newton Trust [18.23(v)].

## Abbreviations

FID: Free Induction Decay
CSI: Chemical Shift Imaging
PPA: phenylphosphonic acid
PCr: Phosphocreatine
ATP: adenosine triphosphate
DPG: 2,3-diphosphoglyerate
PDE: Phosphodiesters
CV: Cine Vascular
WSVD: Whitened Singular Value Decomposition
BISTRO: B_1_-InSensitive TRain to Obliterate signal
AMARES: Advanced Method for Accurate, Robust and Efficient Spectral fitting
AFP: Adiabatic Full Passage
RFPA: Radiofrequency Power Amplifier
CMD: Cardio Microvascular Dysfunction
LVEF: Left Ventricular Ejection Fraction
CS: Circumferential Strain
RS: Radial Strain
AIC: Akaike Information Criterion
PSF: Point Spread Function
BMI: Body Mass Index

## Declaration of generative AI and AI-assisted technologies in the manuscript preparation process

During the preparation of this work, the author(s) used Claude Sonnet 4.6 (Anthropic, PBC, San Francisco, United States) to assist with text writing and MATLAB code optimization for figure generation. The author(s) reviewed and edited the output as needed and take full responsibility for the content of the published article.

## Supplementary Information

**Figure SI1:**
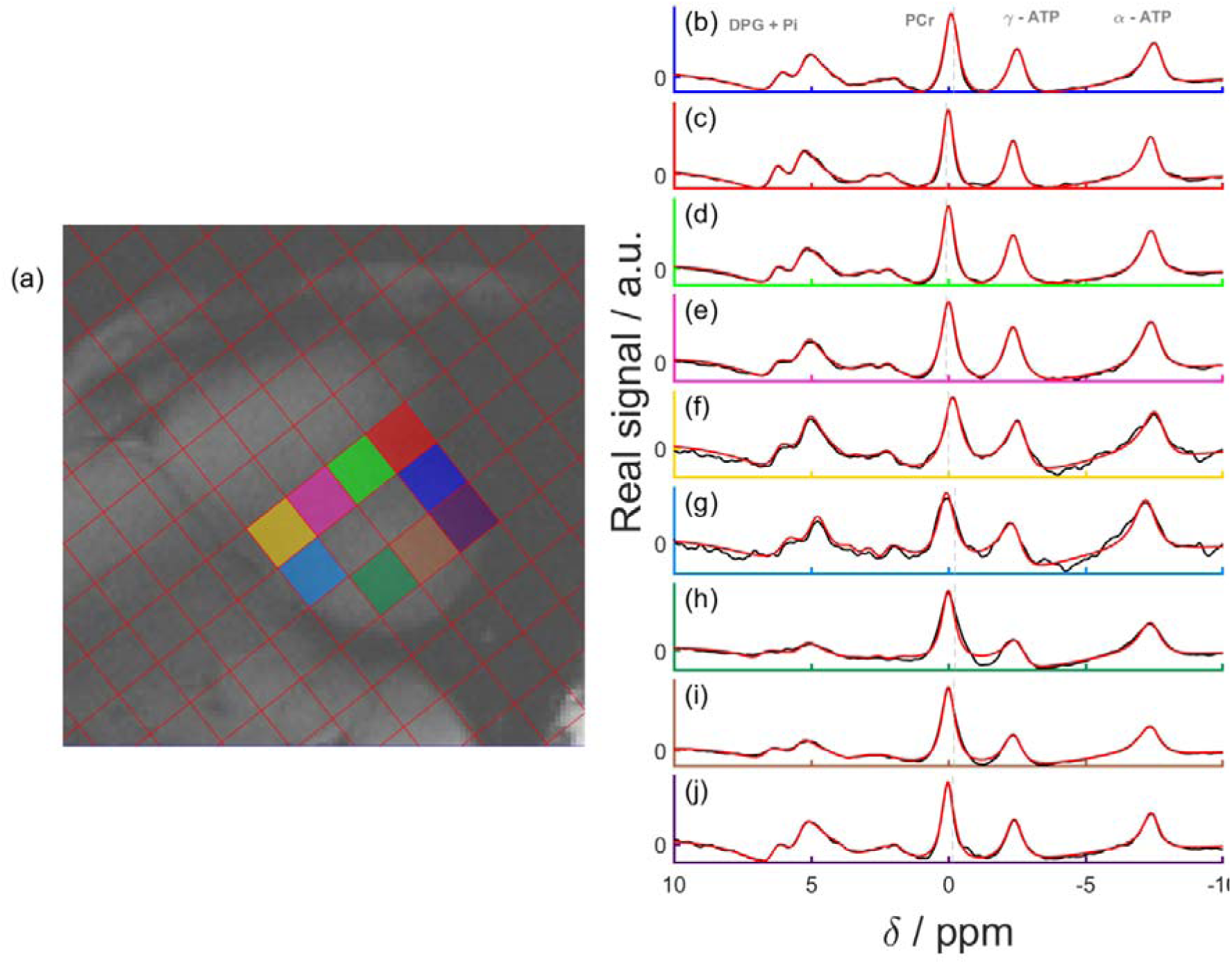
^31^P-MRSI results for a volunteer. Mid-short axis GRE localiser acquired with breath hold with CSI matrix overlaid on the left. Spectra from the corresponding voxels. Both fit (in red) and raw data (in black) are shown.

**Figure SI2:**
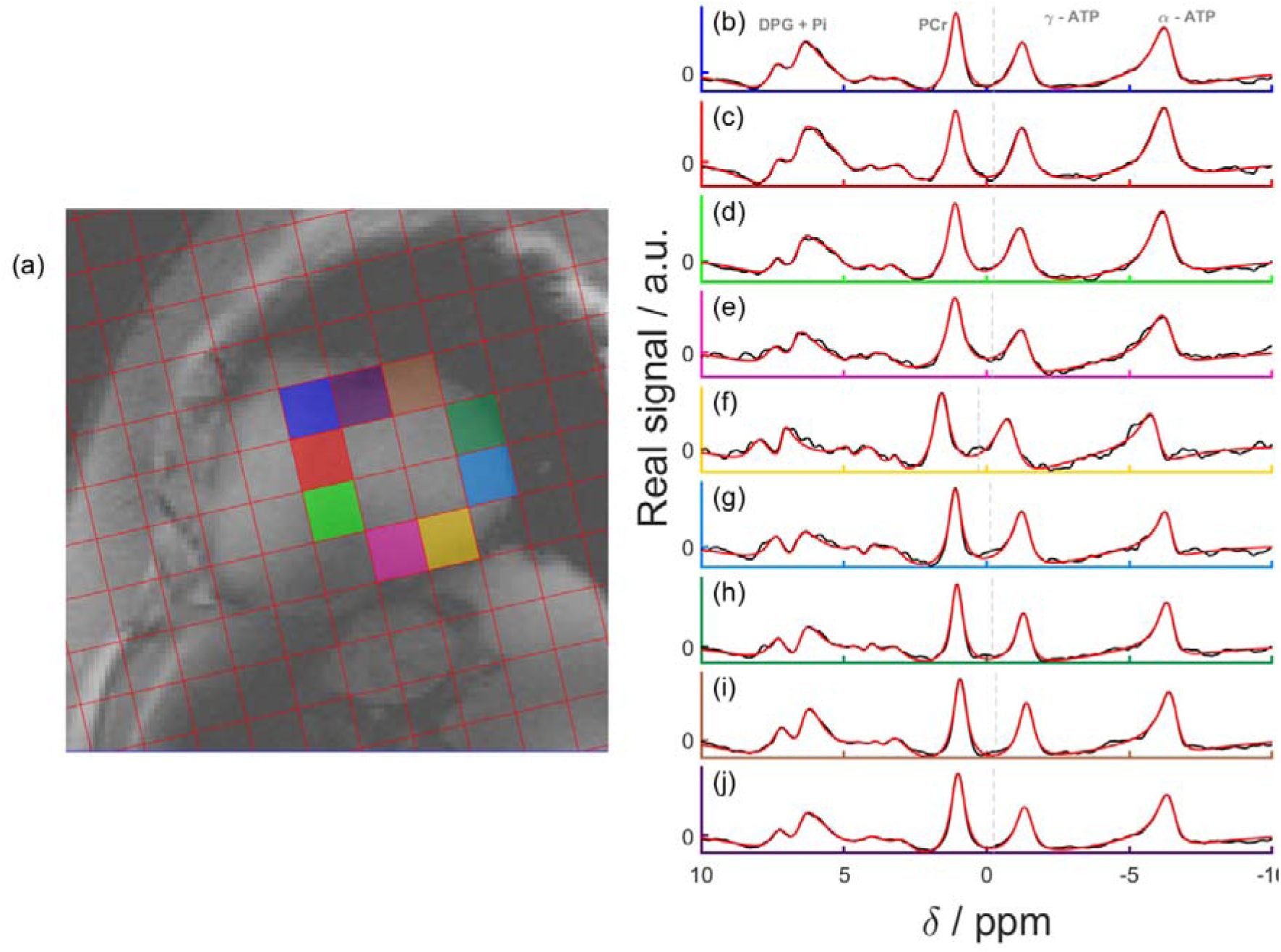
^31^P-MRSI results for a diabetic patient. Mid-short axis GRE localiser acquired with breath hold with CSI matrix overlaid on the left. Spectra from the corresponding voxels. Both fit (in red) and raw data (in black) are shown.

**Figure SI3:**
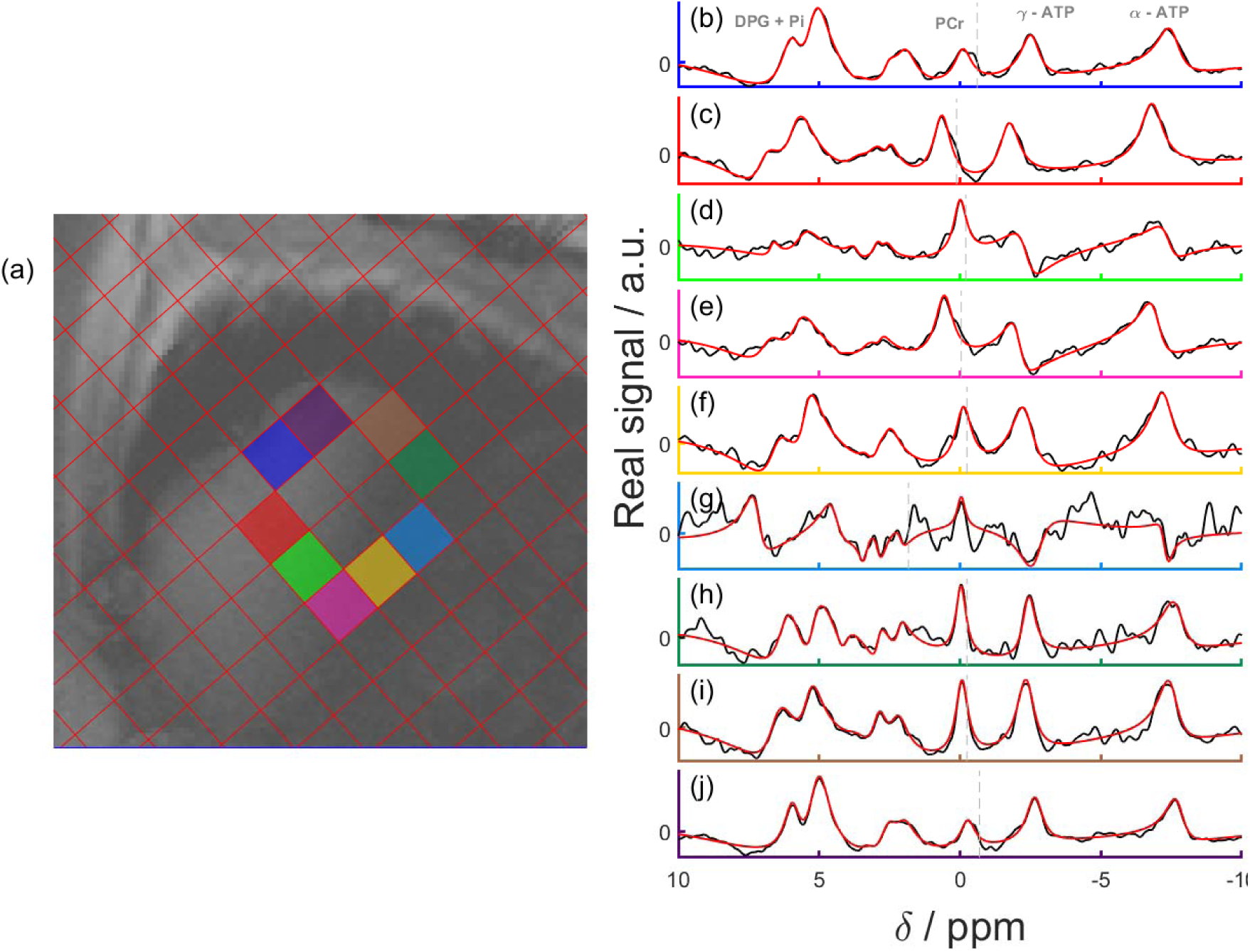
^31^P-MRSI results for a heart failure patient pre-treatment. Mid-short axis GRE localiser acquired with breath hold with CSI matrix overlaid on the left. Spectra from the corresponding voxels. Both fit (in red) and raw data (in black) are shown.

**Figure SI5:**
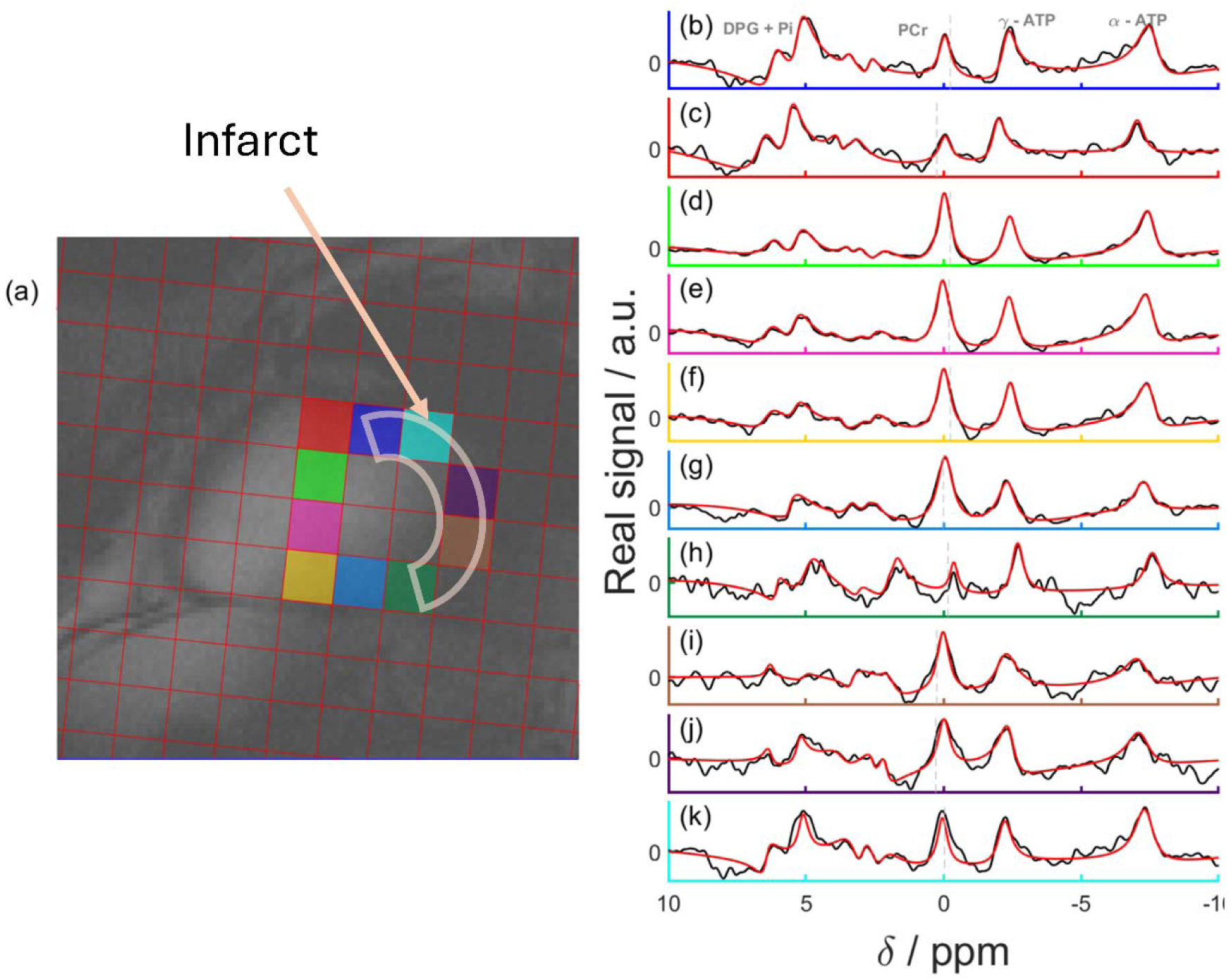
^31^P-MRSI results for a ischaemic patient. Mid-short axis GRE localiser acquired with breath hold with CSI matrix overlaid on the left. Spectra from the corresponding voxels. Both fit (in red) and raw data (in black) are shown.

