## Supplementary code for power calculation for "Regional cardiac ^31^P-MRSI at 7T: detection of spatially heterogeneous patterns of myocardial energetic impairment": PowerCalculations_effectSizes_202608.m

```
%% ===== POWER CALCULATIONS: VERIFICATION =====
% All calculations use two-sided alpha = 0.05
% Reproducibility informed by Karkouri et al. MRM 2025 (ref 23)
% CR_septal = 0.22, CR_lateral = 0.88

alpha      = 0.05;
pow_target = 0.80;

fprintf('========================================\n');
fprintf('POWER CALCULATIONS — VERIFICATION\n');
fprintf('========================================\n\n');

%% --- 1. GROUP-LEVEL KW DIFFERENCES ---
fprintf('--- 1. Group-level KW (eta2 = 0.57, k = 4 groups) ---\n');

eta2_obs = 0.57;
k        = 4;         % Healthy, Diabetic, HF Pre, Ischemic
N_range  = 8:2:100;
power_KW = zeros(size(N_range));

for i = 1:numel(N_range)
    N      = N_range(i);
    lambda = eta2_obs * (N - 1);
    power_KW(i) = 1 - ncx2cdf(chi2inv(1-alpha, k-1), k-1, lambda);
end

N_needed_KW      = N_range(find(power_KW >= pow_target, 1));
n_per_group_KW   = ceil(N_needed_KW / k);

fprintf('N_total needed: %d\n', N_needed_KW);
fprintf('n per group:    %d\n\n', n_per_group_KW);

%% --- 2. PAIRWISE RANK-SUM: HV vs HF (r = 0.96) ---
fprintf('--- 2. Pairwise rank-sum HV vs HF (r = 0.96) ---\n');

r_hf   = 0.96;
d_hf   = 2*r_hf / sqrt(1 - r_hf^2);
n_pair = 3:30;
pow_pair_hf = zeros(size(n_pair));

for i = 1:numel(n_pair)
    n     = n_pair(i);
    df    = 2*n - 2;
    delta = d_hf * sqrt(n/2);
    tcrit = tinv(1 - alpha/2, df);
    pow_pair_hf(i) = 1 - nctcdf(tcrit, df, delta) + nctcdf(-tcrit, df, delta);
end

n_needed_hf = n_pair(find(pow_pair_hf >= pow_target, 1));
fprintf('n per group needed: %d\n\n', n_needed_hf);

%% --- 3. PAIRWISE RANK-SUM: HV vs Ischemic (r = 1.00) ---
fprintf('--- 3. Pairwise rank-sum HV vs Ischemic (r = 1.00) ---\n');
% r = 1.00 means perfect rank separation — minimum n = 3 achieves this
% by definition of the rank-sum test. Report descriptively.
fprintf('r = 1.00: complete rank separation detected at n = 3.\n');
fprintf('Minimum detectable n for rank-sum significance: 3 per group.\n\n');

%% --- 4. SEGMENT-LEVEL SPEARMAN: SEPTAL SEGMENTS ---
fprintf('--- 4. Segment-level Spearman, septal (rs = 0.50-0.56) ---\n');
fprintf('Bonferroni correction for 6 segment comparisons (alpha = %.4f per test)\n\n', ...
    alpha/6);

r_vals   = [0.50, 0.53, 0.56];
n_subj   = 5:5:100;

fprintf('%-8s  %-12s  %-12s\n','rs','n_subjects','n_segments');
fprintf('%s\n', repmat('-',35,1));

for rv = 1:numel(r_vals)
    rs   = r_vals(rv);
    z_r  = 0.5 * log((1+rs)/(1-rs));
    z_crit = norminv(1 - (alpha/6)/2);   % Bonferroni corrected

    pow_seg = zeros(size(n_subj));
    for i = 1:numel(n_subj)
        n_obs    = n_subj(i) * 6;
        se_z     = 1/sqrt(n_obs - 3);
        z_stat   = z_r / se_z;
        pow_seg(i) = 1 - normcdf(z_crit - z_stat) + normcdf(-z_crit - z_stat);
    end

    n_need = n_subj(find(pow_seg >= pow_target, 1));
    if isempty(n_need)
        fprintf('%-8.2f  %-12s  %-12s\n', rs, '>100', '>600');
    else
        fprintf('%-8.2f  %-12d  %-12d\n', rs, n_need, n_need*6);
    end
end

%% --- 5. SEGMENT-LEVEL: ACCOUNTING FOR MEASUREMENT NOISE (CR) ---
fprintf('\n--- 5. Segment-level with measurement noise (CR from ref 23) ---\n');
fprintf('CR_septal = 0.22, mean PCr/ATP_HV = 1.78\n');
fprintf('Measurement SD = CR / (2*sqrt(2)) = %.3f\n', 0.22/(2*sqrt(2)));
fprintf('This attenuates observed rs by factor: signal_var/(signal_var + noise_var)\n\n');

% Attenuation of correlation due to measurement error (reliability correction)
% r_true = r_obs / sqrt(reliability)
% reliability = 1 - (noise_var / total_var)
% For PCr/ATP: signal SD from HV data ~ 0.21, noise SD = CR/(2*sqrt(2))

sigma_signal = 0.21;           % between-subject SD in HV
sigma_noise  = 0.22/(2*sqrt(2));  % from CR_septal
reliability  = sigma_signal^2 / (sigma_signal^2 + sigma_noise^2);
fprintf('Signal SD: %.3f, Noise SD: %.3f\n', sigma_signal, sigma_noise);
fprintf('Reliability coefficient: %.3f\n', reliability);
fprintf('Attenuation factor on observed rs: %.3f\n\n', sqrt(reliability));

% Noise-corrected required n for rs = 0.50 (observed), 
% true rs = observed / sqrt(reliability)
r_obs_vals = [0.50, 0.53, 0.56];
fprintf('%-12s  %-12s  %-14s  %-12s\n', ...
    'rs_obs','rs_true','n_subj (noise-corrected)','n_segments');
fprintf('%s\n', repmat('-',55,1));

for rv = 1:numel(r_obs_vals)
    rs_obs  = r_obs_vals(rv);
    rs_true = min(rs_obs / sqrt(reliability), 0.999);
    z_r     = 0.5 * log((1+rs_true)/(1-rs_true));
    z_crit  = norminv(1 - (alpha/6)/2);

    pow_seg = zeros(size(n_subj));
    for i = 1:numel(n_subj)
        n_obs  = n_subj(i) * 6;
        se_z   = 1/sqrt(n_obs - 3);
        z_stat = z_r / se_z;
        pow_seg(i) = 1 - normcdf(z_crit - z_stat) + normcdf(-z_crit - z_stat);
    end

    n_need = n_subj(find(pow_seg >= pow_target, 1));
    if isempty(n_need)
        fprintf('%-12.2f  %-12.2f  %-24s  %-12s\n', rs_obs, rs_true, '>100', '>600');
    else
        fprintf('%-12.2f  %-12.2f  %-24d  %-12d\n', rs_obs, rs_true, n_need, n_need*6);
    end
end

%% --- 6. LATERAL WALL: HIGHER NOISE ---
fprintf('\n--- 6. Lateral wall with CR = 0.88 ---\n');
sigma_noise_lat  = 0.88/(2*sqrt(2));
reliability_lat  = sigma_signal^2 / (sigma_signal^2 + sigma_noise_lat^2);
fprintf('Lateral noise SD: %.3f\n', sigma_noise_lat);
fprintf('Lateral reliability: %.3f\n', reliability_lat);
fprintf('If true rs = 0.50, observed rs attenuated to: %.3f\n\n', ...
    0.50 * sqrt(reliability_lat));

rs_lat  = 0.50 * sqrt(reliability_lat);
if rs_lat > 0.05
    z_r    = 0.5 * log((1+rs_lat)/(1-rs_lat));
    z_crit = norminv(1 - (alpha/6)/2);
    pow_lat = zeros(size(n_subj));
    for i = 1:numel(n_subj)
        n_obs  = n_subj(i) * 6;
        se_z   = 1/sqrt(n_obs - 3);
        z_stat = z_r / se_z;
        pow_lat(i) = 1 - normcdf(z_crit - z_stat) + normcdf(-z_crit - z_stat);
    end
    n_need_lat = n_subj(find(pow_lat >= pow_target, 1));
    if isempty(n_need_lat)
        fprintf('n subjects needed (lateral, 80%% power): >%d\n', max(n_subj));
    else
        fprintf('n subjects needed (lateral, 80%% power): %d (%d segments)\n', ...
            n_need_lat, n_need_lat*6);
    end
else
    fprintf('Observed rs after attenuation (%.3f) too small to detect.\n', rs_lat);
    fprintf('Motion correction required before lateral wall correlations are detectable.\n');
end

fprintf('\n===== Power calculation verification complete =====\n');
```
